# Genetic risk score for coronary artery calcification predicts coronary artery disease beyond traditional risk factors

**DOI:** 10.1101/2024.03.27.24304983

**Authors:** Pashupati P. Mishra, Binisha H. Mishra, Leo-Pekka Lyytikäinen, Sirkka Goebeler, Mika Martiskainen, Emma Hakamaa, Marcus E. Kleber, Graciela E. Delgado, Winfried März, Mika Kähönen, Pekka J. Karhunen, Terho Lehtimäki

**Affiliations:** Department of Clinical Chemistry, Faculty of Medicine and Health Technology, Tampere University, Tampere, Finland; Finnish Cardiovascular Research Center Tampere, Faculty of Medicine and Health Technology, Tampere University, Tampere, Finland; Department of Clinical Chemistry, Fimlab Laboratories, Tampere, Finland; Forensic Medicine, Finnish Institute for Health and Welfare, Helsinki, Finland; Faculty of Medicine and Health Technology, Tampere University and Fimlab Laboratories, Tampere, Finland; Vth Department of Medicine, University Medicine Mannheim, Medical Faculty Mannheim of the University of Heidelberg, Mannheim, Germany; SYNLAB MVZ Humangenetik Mannheim, Mannheim, Germany; Synlab Academy, SYNLAB Holding Deutschland GmbH, Mannheim and Augsburg, Germany; Department of Clinical Physiology, Tampere University Hospital, Tampere Finland

**Keywords:** Genetic risk score, coronary artery calcification, coronary artery disease, prediction

## Abstract

**Aim:** The modest added predictive value of the existing genetic risk scores (GRSs) for coronary artery disease (CAD) could be partly due to missing genetic components, hidden in the genetic architecture of intermediate phenotypes such as coronary artery calcification (CAC). In this study, we investigated the predictive ability of CAC GRS for CAD.

**Materials and methods:** We investigated the association of CAC GRSs with CAD and coronary calcification among the participants in the Ludwigshafen Risk and Cardiovascular Health study (LURIC) (n=2742), the Tampere Vascular Study (TVS) (n=133), and the Tampere Sudden Death Study (TSDS) (n=660) using summary data from the largest multi-ancestry GWAS meta-analysis of CAC to date. Added predictive value of the CAC GRS over the traditional CVD risk factors was tested with standard train–test machine learning approach using the LURIC data, which had the largest sample size.

**Results:** CAC GRS was significantly associated with CAD in LURIC (OR=1.41, 95% CI [1.28–1.55]), TVS (OR=1.79, 95% CI [1.05–3.21]) as well as in TSDS (OR=4.20, 95% CI [1.74–10.52]). CAC GRS showed strong association with calcification areas in left (OR=1.78, 95% CI [1.16-2.74]) and right (OR=1.71, 95% CI [1.98-2.67]) coronary arteries. In addition, there was statistically significant added predictive value of the CAC GRS for CAD over the used traditional CVD risk factors (AUC 0.734 vs 0.717, p-value = 0.02).

**Conclusions:** This study showed that CAC GRS is a new risk marker for CAD in three European cohorts, with added predictive value over the traditional CVD risk factors.

## 1. Background

Cardiovascular diseases (CVDs) are a key global public health issue with major economic as well as human costs globally [Wilkins et al., 2017]. A major focus on prevention of CVDs is crucial to achieve the United Nations member states’ commitment to a 25% reduction in premature CVD mortality by 2025 [Roth et al., 2015]. Atherosclerosis, the underlying pathology behind most CVDs, begins decades before the clinical manifestations and silently reaches a stage where it can only be slowed down but not be reversed [Hansson, 2005]. Identifying individuals at an early stage of the cardiovascular (CV) cascade who do not yet have symptoms (subclinical atherosclerosis) is of high importance in preventive cardiology.

Genetic risk score (GRS) is a system-level approach that combines single-nucleotide polymorphisms (SNPs) to enhance the predictive ability for a polygenic disease [Knowles et al., 2018]. GRS is an attractive target as a potential predictor because these genetic variants remain stable across the lifespan and can thus provide information on an individual’s disease predisposition from birth and early age to old age. Several genome-wide association studies (GWASs) have characterized the genetic makeup of CVDs, including atherosclerosis-related [Franceschini et al., 2018] and coronary artery disease (CAD)-related SNPs [Nelson et al., 2017]. A GRS calculated with all validated CVD-related SNPs may also predict CVDs in the early stages. However, the SNPs identified so far with traditional GWASs explain only a small fraction (10–20%) of CVD heritability [Nikpay et al., 2015; Nelson et al., 2017; Forgo et al., 2018]. The identification of SNPs explaining the missing heritability is important for a robust GRS and, therefore, constitutes a crucial step forward for the early prediction of CVD. The utility of GRS in CVD risk prediction models is still in the early phase. The modest added predictive value shown by recent studies such as by [Elliott et al., 2020] could partly be due to the missing genetic components hidden within the genetic architecture of intermediate phenotypes of CVD, such as coronary artery calcification (CAC), that are involved in the pathogenesis of CVD. Indeed, although there is shared etiology between CVD and CAC, a recent comprehensive GWAS study of CAC found several new CAC associated genes that have so far not been reported to be associated with CVD [Kavousi et al., 2023]. CAC score, a measure of the amount of calcified plaque in coronary arteries, is a highly reliable indicator of cardiovascular disease beyond the established CVD risk variables [Detrano et al., 2008; Greenland et al., 2018]. According to the American College of Cardiology (ACC)/ American Heart Association (AHA) guidelines, CAC scores are useful in directing treatment for those with intermediate risk of cardiovascular events [Arnett et al., 2019]. However, CAC increases with age and therefore CAC test is not recommended for younger individuals as detectable calcification of the lipid rich plaques takes time. Genetic variants identified with GWAS of CAC might help in developing GRS with CAC equivalent predictive ability for CVD. The ability to predict CVD utilizing CAC GRS can have important ramifications for optimizing lifestyle practices in primary prevention.

The main objective of the current study was to investigate whether GRS for CAC (CAC GRS) is associated with angiographically or morphometrically verified CAD in three European cohorts. The CAC GRS was calculated using genetic summary statistics from the most recent GWAS study of CAC [Kavousi et al., 2023]. We aimed to confirm the link between CAC associated SNPs and calcification by studying the association of CAC GRS with calcification areas measured in autopsy coronary arteries. In addition to the association study of the CAC GRS with CAD, we also assessed its added predictive value for CAD over the traditional CVD risk factors (age, sex, total cholesterol, HDL cholesterol, systolic blood pressure, treatment for hypertension and smoking habit) that are included in the Framingham risk score (FRS) [D’Agostino et al., 2008].

## 2. Methods

### 2.1 Study participants

This study was based on the Ludwigshafen Risk and Cardiovascular Health (LURIC) study participants as discovery cohort and the Tampere Vascular Study (TVS) participants and the Tampere Sudden Death Study (TSDS) as validation cohorts. The LURIC study comprises of 3,316 European patients referred to the Cardiac Center Ludwigshafen (Germany) for invasive coronary angiography between July 1997 and January 2000 [Winkelmann et al., 2001]. Clinical indications for angiography were chest pain or a positive non-invasive stress test suggestive of myocardial ischemia. To limit clinical heterogeneity, individuals suffering from acute illnesses other than acute coronary syndrome (ACS), chronic non-cardiac diseases and a history of malignancy within the five past years were excluded. The study protocol was approved by the ethics committee of the ‘Landesärztekammer Rheinland-Pfalz’ [#837.255.97(1394)]. The present study is based on 2742 patients (women: 29%), aged 17–92 years, who had genotype data as well as CAD and traditional CVD risk factors data used in this study. The patients with 50% or more stenosis in one of the major coronary arteries were defined as CAD cases.

Tampere Vascular Study (TVS) constitutes individuals at high risk of cardiovascular diseases, events and deaths who have undergone an exercise stress test at Tampere University Hospital [Nieminen et al., 2006]. This study was based on genotype data from a subpopulation of 133 individuals with 98 angiographically verified cases of CAD followed-up during 2008. The patients with at least 50% stenosis in either left main coronary artery, left anterior descending (LAD) coronary artery or right coronary artery were defined as CAD cases. The participants were 38–80 years old and 29% of them were women. The study has been approved by the Ethics Committee of Tampere Hospital District. All studies were conducted according to the declaration of Helsinki, with the informed consent from individual patient involved.

The Tampere Sudden Death Study (TSDS) consists of cross-sectional population autopsy samples of deaths occurring out of hospital for any reason. This study involved 660 cases who had both genetic data and coronary measurements available. Computer-assisted morphometry (Olympus Cell-D software) was used to measure the percentage area of calcified lesions from the five centimeters long piece of the proximal part of the LAD and right coronary arteries (RCA). The stenosis percentage was measured from microscopic sections of the coronary arteries. TSDS cases with 50% or more stenosis in either LAD coronary artery including left main part of the artery or RCA were defined as CAD cases. The participants were aged 18–94 years and 28% of them were women. The CVD risk factor data such as smoking and hypertension were collected by interviewing a spouse, a relative, or a close friend of the deceased. The permission to collect the data was obtained from the ethical committee of Tampere University hospital (Permission number R09097) and from the National Supervisory Authority for Welfare and Health.

### 2.2 Traditional CVD risk factor measurements

Traditional risk factors for CVD included in this study were age, sex, total cholesterol, high density lipoprotein (HDL) cholesterol, smoking habit, systolic blood pressure and hypertension treatment. The assay methods have been reported in detail previously for the LURIC participants [Winkelmann et al., 2001] and for the TVS participants [Nieminen et al., 2006]. The selection of the risk factors was based on the six coronary risk factors used in the Framingham risk score [D’Agostino et al., 2008]. Only age, sex, and body mass index (BMI) were available CVD risk factors in the TSDS, whereas data on smoking habit and hypertension were gathered via postal questionnaire from spouses or close relatives and were available for 228 (34.5%) out of the 660 participants.

### 2.3 Genotyping, quality control and genotype imputation

Genomic DNA for the LURIC study participants was prepared from EDTA anticoagulated peripheral blood by means of a common salting-out procedure. The Affymetrix Genome-Wide Human SNP Array 6.0 was used for genotyping. Genomic DNA for the TVS was extracted from peripheral blood leukocytes using QIAamp DNA Blood Minikit and automated biorobot M48 extraction (Qiagen, Hilden, Germany). Genotyping was done using the Illumina HumanHap660W-Quad BeadChip according to the manufacturer’s recommendation. In TSDS, DNA was extracted from postmortem blood using a Qiagen DNA extraction kit. Illumina HumanCoreExome-12 were used as genotyping platform and genotypes were called with Illumina GenCall algorithm. The quality control filters applied to the LURIC genotype data included: sample call rate < 0.95, SNP call rate < 0.98, Hardy-Weinberg equilibrium test p-value < 1 x 10^-04^, and minor allele frequency < 0.01. Genotype imputation was completed with the Haplotype Reference Consortium (HRC) imputation panel as a reference. The quality control filters applied for the TVS and TSDS included: sample and SNP call rate <0.95, Hardy-Weinberg equilibrium test p-value<10^-6^, and minor allele frequency < 5%. Genotype imputation in both TVS and TSDS was performed using SNPTEST v2.3.0 and 1000 Genomes phase 1 version 3 haplotypes as reference. All the 11 independent SNPs used in the construction of CAC GRS in all the three cohorts had a squared correlation > 0.40 between imputed and true genotypes.

### 2.4 Biostatistical analysis

All biostatistical analyses and data processing were performed using the statistical package R version 4.1.0 [R Core Team, 2020]. The CAC GRSs in all the three cohorts (LURIC, TVS and TSDS) were calculated using GWAS summary statistics of 11 independent SNPs associated with CAC [Kavousi et al., 2023] as sum of effect allele dosages or counts of the 11 SNPs weighted by their corresponding effect sizes, as estimated from the GWAS study [Kavousi et al., 2023]. The CAC GRS was standardized to have zero mean and unit standard deviation. Association of the CAC GRS with CAD in the LURIC, TVS and TSDS participants was tested using two logistic regression models, one adjusted for age, sex and BMI (model 1) and the other adjusted for traditional coronary risk factors used in the FRS, namely age, sex, smoking status, HDL cholesterol, total cholesterol, systolic blood pressure and treatment for hypertension [D’Agostino et al., 2008] (model 2). Additionally, the model 2 in LURIC and TVS was also adjusted for the first five principal components of the genetic data to adjust for potential bias due to population structure and statin usage. Model 2 in TSDS was adjusted for age, sex, BMI, smoking, hypertension and the first five principal components of the genetic data. Measurement data on clinical lipids, blood pressure and statin usage were not available among the TSDS participants. Association of CAC GRS with calcified plaque percentage in LAD as well as RCA of the TSDS participants was tested using beta regression models using *betareg* R package [Cribari-Neto & Zeileis, 2010]. The analyses were adjusted for age, sex, BMI, smoking, hypertension and the first five principal components of the genetic data.

Added predictive ability of the CAC GRS over the traditional CVD risk factors was assessed using the LURIC data. A reference prediction model for CAD including only the traditional CVD risk factors as predictors was built. Then, the added predictive value of the CAC GRS for CAD was accessed by comparing logistic regression-based prediction model including the traditional CVD risk factors and CAC GRS (test prediction model) with the reference prediction model as described elsewhere [Mishra et al., 2021]. Assessment of the predictive value of the CAC GRS was performed using a standard train-test split machine learning approach, i) fitting models to training data (70% data), ii) testing the models on test data (30% data), and iii) calculating the area under the receiver operating curve (AUC) for assessment of the predictive model. The variance of AUC was estimated by repeating the model fitting and validation for 1000 bootstraps of the original data. Added predictive ability of the CAC GRS over the traditional CVD risk factors was tested by taking differences between AUCs (ΔAUC) obtained from the reference and test prediction models over 1000 bootstraps of the original data. The statistical significance of the difference (p-value) was estimated by counting the proportion of ΔAUC less than zero.

## 3. Results

### 3.1 Study population characteristics

Clinical characteristics of the study participants from the LURIC, TVS and TSDS cohorts are presented in Table 1. A total of 2742 individuals had complete data on genetics, angiographically verified CAD and traditional CVD risk factors in LURIC. In TVS, 133 participants had complete data on genetics, angiographically verified CAD and traditional CVD risk factors. In case of TSDS, there were 622 participants with complete data on genetics, morphometrically verified CAD, calcified plaque area percentage in the LAD and RCA, age, sex and BMI. Data on smoking habit and medication for hypertension was available for 228 out of the 622 TSDS participants. Data from all the three cohorts, stratified by case–control, is presented in Table 1

**Table 1.** Clinical characteristics of the study cohorts.

|  | LURIC |  |  | TVS |  |  | TSDS |  |  |
| --- | --- | --- | --- | --- | --- | --- | --- | --- | --- |
| Variables | CAD Cases | Controls | P-value | CAD Cases | Controls | P-value | CAD Cases | Controls | P-value |
| Number of subjects (%) | 1912 (70) | 830 (30) | - | 98 (74) | 35 (26) | - | 327 (53) | 295 (47) | - |
| Female (%) | 425 (22) | 367 (44) | - | 28 (29) | 10 (29) | - | 79 (24) | 74 (25) | - |
| Age, years | 64 (29-92) | 60 (17-88) | $8.8 \times 10^{-16}$ | 60 (39-80) | 56 (38-72) | 0.01 | 69 (24-95) | 58 (17-95) | $<2.2 \times 10^{-16}$ |
| Body mass index (kg/m<sup>2</sup>) | 27.4 (16.3-46.2) | 27.6 (16.7-46.1) | 0.3 | 27.8 (18.9-41.3) | 27.5 (18.7-36.9) | 0.8 | 28.7 (16.4-54.1) | 28.9 (14.6-64.9) | 0.7 |
| Systolic blood pressure (mmHg) | 142.3 (76.7-230) | 138.6 (73.7-228.3) | $7.2 \times 10^{-05}$ | 134.9 (86-192) | 135.1 (104-176) | 0.97 | - | - | - |
| Total cholesterol (mg/dL) | 205.2 (100-442) | 215.2 (79-453) | $2.4 \times 10^{-08}$ | 94.1 (55.8-135) | 100 (72.0-151.2) | 0.2 | - | - | - |
| HDL cholesterol (mg/dL) | 37.1 (2-104) | 41.5 (15-91) | $<2.2 \times 10^{-16}$ | 25.4 (12.2-55.6) | 26.6 (13.9-38.9) | 0.4 | - | - | - |
| LDL cholesterol (mg/dL) | 114.4 (20-329) | 120.9 (21-361) | $4.7 \times 10^{-06}$ | 56.8 (19.8-95.4) | 62.0 (36-99) | 0.09 | - | - | - |
| Smoking (%) | 1337 (70) | 423 (51) | $<2.2 \times 10^{-16}$ | 61 (62) | 21 (60) | 0.97 | 68/130 (52) | 56/98 (57) | 0.64 |
| Medication for hypertension | 1185 (62) | 431 (52) | $1.1 \times 10^{-06}$ | 93 (95) | 28 (80) | 0.02 | 88/130 (68) | 52/98 (53) | 0.008 |
| Statin usage | 1138 (60) | 176 (21) | $<2.2 \times 10^{-16}$ | 81 (83) | 11 (31) | $6 \times 10^{-08}$ | - | - | - |
| Stenosis in LAD (%) | - | - | - | - | - | - | 61.5 (5-100) | 24.7 (0-49.3) | $<2.2 \times 10^{-16}$ |
| Stenosis in RCA (%) | - | - | - | - | - | - | 52.5 (0-100) | 18.4 (0-49.2) | $<2.2 \times 10^{-16}$ |
| Calcium in LAD (%) | - | - | - | - | - | - | 28.9 (0-100) | 6.3 (0-79.1) | $<2.2 \times 10^{-16}$ |
| Calcium in RCA (%) | - | - | - | - | - | - | 25.7<br>(0 - 100) | 4.3<br>(0-88.8) | <2.2 x 10<sup>-16</sup> |

*Definitions:* Patients with 50% or more stenosis in one of the major coronary arteries were defined as CAD cases in both LURIC and TVS cohorts. Autopsy cases with 50% or more stenosis in either left anterior descending (LAD) coronary artery including left main part of the artery or right coronary artery (RCA) were defined as CAD cases in the TSDS cohort.
*Abbreviations:* LURIC, the Ludwigshafen Risk and Cardiovascular Health Study; TVS, the Tampere Vascular Study; TSDS, the Tampere Sudden Death Study; CAD, Coronary Artery Disease; LAD, Left anterior descending.

### 3.2 Association of CAC GRS with CAD in LURIC, TVS and TSDS cohorts

In LURIC, CAC GRS was associated with angiographically verified CAD in both model 1 (OR of 1.45, 95% CI [1.33–1.59]) and model 2 (OR=1.41, 95% CI [1.28–1.55]) [Table 2]. Among the traditional CVD risk factors, age, sex, HDL cholesterol, smoking habit and medication for hypertension were associated with CAD with statistical significance of p-value < 0.05. In TVS, CAC GRS was associated with angiographically verified CAD in both model 1 (OR=1.64, 95% CI [1.08–2.56]) and model 2 (OR=1.79, 95% CI [1.05–3.21]). Besides the CAC GRS, age, sex, total cholesterol, smoking habit and medication for hypertension were associated with CAD with statistical significance of p-value < 0.05. Similarly, in TSDS, CAC GRS was associated with CAD in both model 1 (OR=2.96, 95% CI [1.79–4.97]) and model 2 (OR=4.20, 95% CI [1.74– 10.52]). Among the traditional risk factors included in the models, only age was associated with CAD with statistical significance (p-value=1.2 x 10^-05^).

**Table 2.** Association of the genetic risk score for coronary artery calcification (CAC GRS) and clinical cardiovascular disease (CVD) risk factors with coronary artery disease (CAD) in the Ludwigshafen Risk and Cardiovascular Health (LURIC) study, Tampere Vascular Study (TVS) and Tampere Sudden Death Study (TSDS) participants.

| Risk factors | LURIC |  |  |  | TVS |  |  |  | TSDS |  |  |  |
| --- | --- | --- | --- | --- | --- | --- | --- | --- | --- | --- | --- | --- |
|  | Model 1 |  | Model 2 |  | Model 1 |  | Model 2 |  | Model 1 |  | Model 2 |  |
|  | OR (95% CI) | P-value | OR (95% CI) | P-value | OR (95% CI) | P-value | OR (95% CI) | P-value | OR (95% CI) | P-value | OR (95% CI) | P-value |
| Age | 1.05<br>(1.04–1.05) | < 2 x 10<sup>-16</sup> | 1.05<br>(1.04 – 1.06) | < 2 x 10<sup>-16</sup> | 1.06<br>(1.01–1.11) | 0.02 | 1.07<br>(1.01–1.15) | 0.04 | 1.05<br>(1.04–1.07) | 3.9 x 10<sup>-16</sup> | 1.06<br>(1.03–1.08) | 1.2 x 10<sup>-5</sup> |
| Sex (male) | 3.49<br>(2.90–4.21) | < 2 x 10<sup>-16</sup> | 2.70<br>(2.16 – 3.39) | < 2 x 10<sup>-16</sup> | 0.86<br>(0.34–2.10) | 0.75 | 4.94<br>(1.26–23.62) | 0.03 | 1.52<br>(1.01–2.28) | 0.04 | 1.29<br>(0.65–2.57) | 0.5 |
| Body mass index (BMI) | 0.99<br>(0.97–1.01) | 0.26 | - | - | 1.03<br>(0.94–1.13) | 0.59 | - | - | 1.01<br>(0.98–1.04) | 0.5 | 1.02<br>(0.97–1.07) | 0.5 |
| Systolic blood pressure | - | - | 1.00<br>(1.00 – 1.01) | 0.19 | - | - | 1.01<br>(0.98–1.04) | 0.51 | - | - | - | - |
| Total cholesterol | - | - | 1.00<br>(0.99 – 1.00) | 0.32 | - | - | 0.48<br>(0.25–0.85) | 0.02 | - | - | - | - |
| HDL cholesterol | - | - | 0.98<br>(0.97 – 0.99) | 5.9 x 10<sup>-07</sup> | - | - | 0.84<br>(0.16–4.57) | 0.83 | - | - | - | - |
| Smoking habit (yes) | - | - | 1.83<br>(1.48 – 2.26) | 1.7 x 10<sup>-08</sup> | - | - | 3.46<br>(1.07–12.24) | 0.04 | - | - | 1.87<br>(0.97–3.71) | 0.06 |
| Medication for hypertension (yes) | - | - | 1.32<br>(1.07 – 1.63) | 0.008 | - | - | 6.79<br>(1.26–41.1) | 0.03 | - | - | 1.29<br>(0.67–2.47) | 0.4 |
| CAC GRS | 1.45<br>(1.33–1.59) | < 2 x 10<sup>-16</sup> | 1.41<br>(1.28 – 1.55) | 3.6 x 10<sup>-12</sup> | 1.64<br>(1.08–2.56) | 0.02 | 1.79<br>(1.05–3.21) | 0.04 | 2.96<br>(1.79–4.97) | 3.1 x 10<sup>-05</sup> | 4.20<br>(1.74–10.52) | 0.002 |

*Abbreviations:* OR, odd ratio; CI, confidence interval.

### 3.3 Association of CAC GRS with calcified plaque area percentage in TSDS participants

We assessed association of the CAC GRS with calcified plaque area percentage in the LAD and RCA of the TSDS participants. There was statistically significant association between the CAC GRC and calcified plaque area in both LAD (OR=1.78, 95% CI [1.16–2.74]) and RCA (OR=1.71, 95% CI [1.09–2.67]) after adjusting for age, sex, BMI, smoking habit, hypertension and the first five principal components of the genetic data [Table 3]. Thus, CAC GRS is a significant predictor of coronary calcification along with age, sex and hypertension.

**Table 3.** Association of the genetic risk score for coronary artery calcification (CAC GRS) with calcified plaque area percentage in the left anterior descending (LAD) and right coronary arteries (RCA).

| Risk factors | Calcified plaque area percentage in the LAD |  | Calcified plaque area percentage in the RCA |  |
| --- | --- | --- | --- | --- |
|  | OR (95% CI) | P-value | OR (95% CI) | P-value |
| Age | 1.03 (1.02–1.04) | $4.7 \times 10^{-9}$ | 1.02 (1.01–1.04) | $1.6 \times 10^{-5}$ |
| Sex (male) | 1.53 (1.09–2.13) | 0.01 | 1.35 (0.96–1.90) | 0.08 |
| Body mass index (BMI) | 0.99 (0.97–1.01) | 0.4 | 1.00 (0.98–1.03) | 0.7 |
| Smoking habit (yes) | 1.06 (0.77–1.46) | 0.7 | 1.09 (0.79–1.52) | 0.6 |
| Hypertension (yes) | 1.62 (1.17–2.24) | 0.004 | 1.53 (1.10–2.15) | 0.01 |
| CAC GRS | 1.78 (1.16–2.74) | 0.009 | 1.71 (1.09–2.67) | 0.02 |

*Abbreviations:* OR, odd ratio; CI, confidence interval.

### 3.4 Assessment of added predictive value of CAC GRS over traditional CVD risk factors

There was statistically significant (AUC 0.734 vs 0.717, p-value = 0.02) added predictive value of the CAC GRS over the traditional CVD risk factors used in this study [Figure 1]. Assessment of the statistical significance was done by calculating the proportion of differences in the AUC (ΔAUC) less than zero obtained from predictive model for CAD diagnosis in the LURIC participants using the traditional CVD risk factors without and with the addition of CAC GRS over 1000 bootstraps.

**Figure 1.**
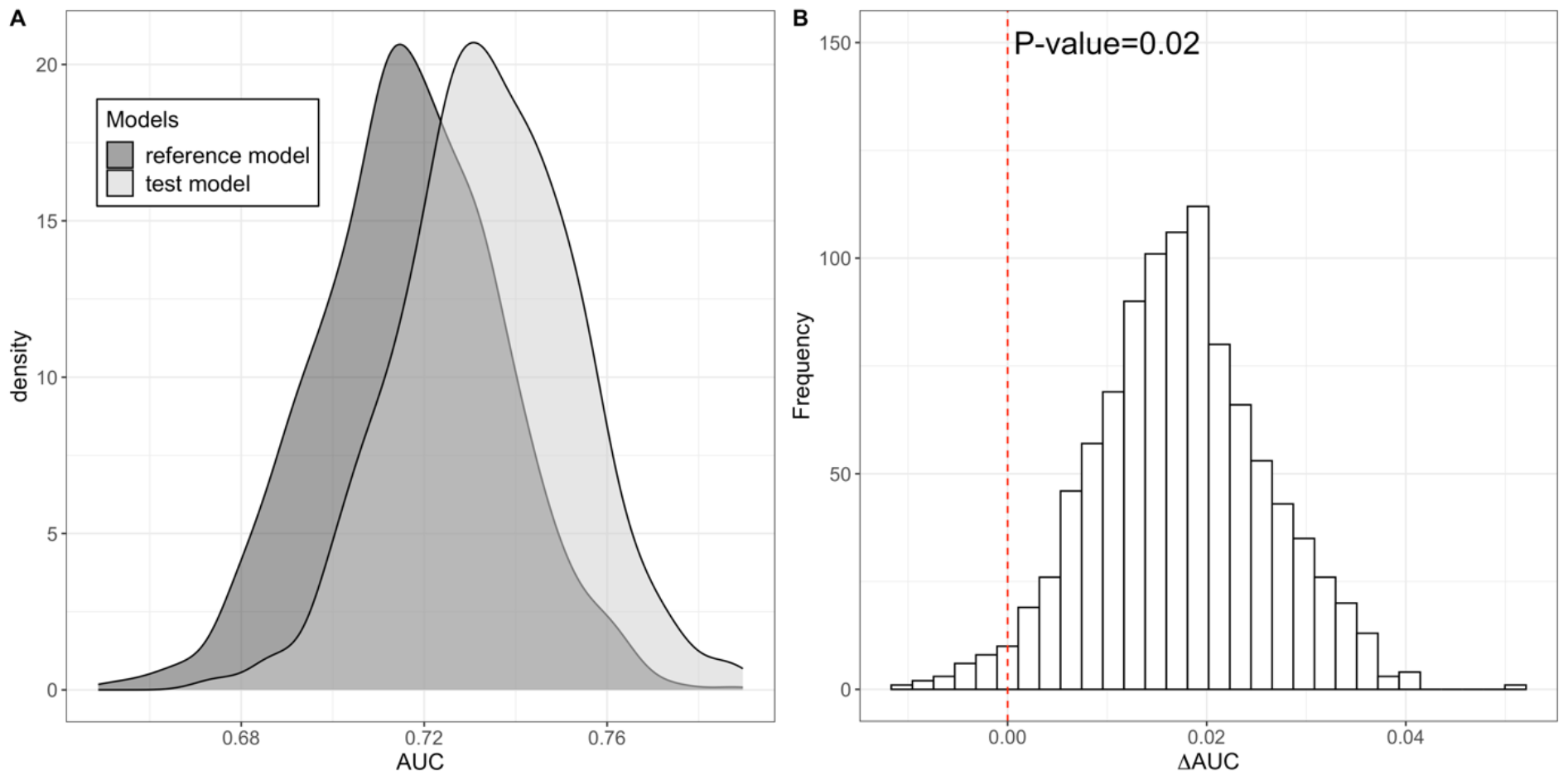
**A**. Density plot illustrating distributions of the area under the receiver operating curve (AUC) obtained from predictive model for coronary artery disease (CAD) diagnosed in the Ludwigshafen Risk and Cardiovascular Health (LURIC) study participants using traditional CVD risk factors without (reference model) and with the addition of genetic risk scores for coronary artery calcification (CAC GRS) (test model) over 1000 bootstraps. **B**. Distribution of differences in the area under the receiver operating curve (ΔAUC) obtained from predictive model for coronary artery disease (CAD) diagnosed in the Ludwigshafen Risk and Cardiovascular Health (LURIC) study participants using traditional CVD risk factors without and with the addition of genetic risk scores for coronary artery calcification (CAC GRS) over 1000 bootstraps. (Test model: *CAD ∼ CAC GRS + age + sex + systolic blood pressure + total cholesterol + HDL cholesterol + smoking habit + medication for hypertension*. Reference model: *CAD ∼ age + sex + systolic blood pressure + total cholesterol + HDL cholesterol + smoking habit + medication for hypertension*.)

## 4. Discussion

We found statistically significant associations of CAC GRS with CAD in all the three European cohorts studied in this study. We also demonstrated that CAC GRS has statistically significant added predictive value for CAD over the traditionally used CVD risk factors. The findings suggest that the genetic variants regulating coronary artery calcification may be used as a tool for CVD risk prediction. Moreover, we found that CAC GRS is significantly associated with morphometrically measured coronary calcification areas, which confirms the earlier findings on the association between CAC related genes and coronary calcification measured with computed tomography at the vessel wall level [Kavousi et al., 2023].

There is consistent evidence suggesting that a predictive value of GRS for CVDs may have clinical utility [Jack et al., 2023]. Efforts to develop or improve existing GRS for CVD risk prediction is important because a robust GRS with high predictability has a potential to shift the paradigm in prevention approaches of CVD. Optimization of the GRS for CVD prediction is crucial to achieve the aimed progress in primordial prevention of CVD [Lloyd-Jones et al., 2010].

The underlying hypothesis of this study was that a part of the missing heritability of CVD might be explained by SNPs associated with intermediate phenotypes of CVD such as CAC [Blanco-Gómez et al., 2016]. CAC is a highly predictive CVD risk marker in asymptomatic individuals that can provide additional predictive value over the traditional CVD risk factors [Detrano et al., 2008; Greenland et al., 2018]. Investigation of the genetic architecture of CAC may reveal SNPs that are critical to understanding the genetic basis of CVD. In this study, we explored whether SNPs associated with CAC, when aggregated as a CAC GRS, are associated with CAD. The findings of this study suggest that CAC GRS might be a useful CVD risk marker with statistically significant added predictive value over the traditionally used risk factors and coronary calcified plaque area. An important benefit of CAC GRS over CAC is that the former is stable throughout the lifespan and therefore can play impactful role in primary prevention of CVD.

This study has some limitations. Some of the traditional CVD risk factors such as systolic blood pressure, HDL cholesterol, total cholesterol and information about statin usage were not available for the autopsy based TSDS participants. Furthermore, due to missing response rate characteristic of postal surveys, data on smoking habit and medication for hypertension was available for only one third of the participants. Also, all the three cohorts included in this study are of European origin, and therefore, studies with populations of different ethnicities are needed.

Based on the results from three different European cohorts, this study showed that CAC GRS is a new risk marker for CAD. The CAC GRS had statistically significant added predictive value over the traditional CVD risk factors and, therefore, can play a crucial role in primary prevention of CVD.

## Data Availability

All data produced in the present study are available upon reasonable request to the authors.

## Conflict of interest

The authors declare that they have no known competing financial interests or personal relationships that could have appeared to influence the work reported in this paper.

### Financial support

This study was supported by the Academy of Finland (Grant number: 349708 for P.P.M and 322098 for T.L). Competitive State Research Financing of the Expert Responsibility area of Tampere University Hospital (grants X51001 and X51401); Juho Vainio Foundation; Finnish Foundation for Cardiovascular Research; Tampere Tuberculosis Foundation; Emil Aaltonen Foundation; Yrjö Jahnsson Foundation; Signe and Ane Gyllenberg Foundation; Diabetes Research Foundation of Finnish Diabetes Association; EU Horizon 2020 (grant 755320 for TAXINOMISIS and grant 848146 for To Aition); Tampere University Hospital Supporting Foundation, Finnish Society of Clinical Chemistry and Jane and Aatos Erkko Foundation. LURIC was supported by the 7th Framework Programs Atheroremo (Grant Agreement number 201668) and RiskyCAD (grant agreement number 305739) of the European Union and by the H2020 Programs TO_AITION (grant agreement number 848146) and TIMELY (grant agreement number 101017424) of the European Union.

### CRediT authorship contribution statement

**Pashupati P. Mishra**: Conceptualization, Methodology, Formal analysis, Investigation, Writing - Original Draft. **Binisha H. Mishra**: Formal analysis, Writing – Review & Editing. **Leo-Pekka Lyytikäinen**: Formal analysis, Writing – Review & Editing. **Sirkka Goebeler**: Resources. **Mika Martiskainen**: Resources. **Emma Hakamaa**: Investigation, Writing – Review & Editing. **Marcus E. Kleber**: Formal analysis, Writing – Review & Editing. **Graciela E. Delgado**: Writing – Review & Editing. **Winfried März**: Resources, Writing – Review & Editing. **Mika Kähönen**: Resources, Writing – Review & Editing. **Pekka J. Karhunen**: Resources, Writing – Review & Editing. **Terho Lehtimäki**: Resources, Writing – Review & Editing.

## References

1. Wilkins, E., Wilson, L., Wickramasinghe, K., & Bhatnagar, P. (2017). European Cardiovascular Disease Statistics 2017. European Heart Network, 94–100. Retrieved from https://www.ehnheart.org

2. Roth, G. A., Nguyen, G., Forouzanfar, M. H., Mokdad, A. H., Naghavi, M., & Murray, C. J. L. (2015). Estimates of global and regional premature cardiovascular mortality in 2025. Circulation, 132(13), 1270–1271. 10.1161/CIRCULATIONAHA.115.016021

3. Hansson, G. K. (2005). Inflammation, Atherosclerosis, and Coronary Artery Disease. New England Journal of Medicine, 352(16), 1685–1695. 10.1056/nejmra043430

4. Knowles, J. W., & Ashley, E. A. (2018). Cardiovascular disease: The rise of the genetic risk score. PLoS Medicine, 15(3). 10.1371/journal.pmed.1002546

5. Franceschini, N., Giambartolomei, C., de Vries, P. S., Finan, C., Bis, J. C., Huntley, R. P., … O’Donnell, C. J. (2018). GWAS and colocalization analyses implicate carotid intima-media thickness and carotid plaque loci in cardiovascular outcomes. Nature Communications, 9(1). 10.1038/s41467-018-07340-5

6. Nelson, C. P., Goel, A., Butterworth, A. S., Kanoni, S., Webb, T. R., Marouli, E., … Deloukas, P. (2017). Association analyses based on false discovery rate implicate new loci for coronary artery disease. Nature Genetics, 49(9), 1385–1391. 10.1038/ng.3913

7. Nikpay, M., Goel, A., Won, H. H., Hall, L. M., Willenborg, C., Kanoni, S., … Farrall, M. (2015). A comprehensive 1000 Genomes-based genome-wide association meta-analysis of coronary artery disease. Nature Genetics, 47(10), 1121–1130. 10.1038/ng.3396

8. Forgo, B., Medda, E., Hernyes, A., Szalontai, L., Tarnoki, D. L., & Tarnoki, A. D. (2018, October 1). Carotid artery atherosclerosis: A review on heritability and genetics. Twin Research and Human Genetics. Cambridge University Press. 10.1017/thg.2018.45

9. Elliott, J., Bodinier, B., Bond, T. A., Chadeau-Hyam, M., Evangelou, E., Moons, K. G. M., … Tzoulaki, I. (2020). Predictive Accuracy of a Polygenic Risk Score-Enhanced Prediction Model vs a Clinical Risk Score for Coronary Artery Disease. JAMA - Journal of the American Medical Association, 323(7), 636–645. 10.1001/jama.2019.22241

10. Kavousi, M., Bos, M. M., Barnes, H. J., Lino Cardenas, C. L., Wong, D., Lu, H., … Miller, C. L. (2023). Multi-ancestry genome-wide study identifies effector genes and druggable pathways for coronary artery calcification. Nature Genetics, 55(10), 1651–1664. 10.1038/s41588-023-01518-4

11. D’Agostino, R. B., Vasan, R. S., Pencina, M. J., Wolf, P. A., Cobain, M., Massaro, J. M., & Kannel, W. B. (2008). General Cardiovascular Risk Profile for Use in Primary Care. Circulation, 117(6), 743–753. 10.1161/circulationaha.107.699579

12. Winkelmann, B. R., März, W., Boehm, B. O., Zotz, R., Hager, J., Hellstern, P., & Senges, J. (2001). Rationale and design of the LURIC study - A resource for functional genomics, pharmacogenomics and long-term prognosis of cardiovascular disease. Pharmacogenomics, 2(1 SUPPL. 1). 10.1517/14622416.2.1.s1

13. Team, R. C. (2021). R: A Language and Environment for Statistical Computing. R Foundation for Statistical Computing. Retrieved from http://www.r-project.org/

14. Mishra, P. P., Mishra, B. H., Lyytikäinen, L. P., Hilvo, M., Juonala, M., Kähönen, M., … Lehtimäki, T. (2021). Assessment of plasma ceramides as predictor for subclinical atherosclerosis. Atherosclerosis Plus, 45, 25–31. 10.1016/j.athplu.2021.09.005

15. Jack W O’Sullivan, Euan A Ashley, Perry M Elliott, Polygenic risk scores for the prediction of cardiometabolic disease, European Heart Journal, Volume 44, Issue 2, 7 January 2023, Pages 89–99, 10.1093/eurheartj/ehac648

16. Lloyd-Jones, D., Adams, R. J., Brown, T. M., Carnethon, M., Dai, S., De Simone, G., … Wylie-Rosett, J. (2010, February). Executive summary: Heart disease and stroke statistics-2010 update: A report from the american heart association. Circulation. 10.1161/CIRCULATIONAHA.109.192667

17. Abraham, G., Malik, R., Yonova-Doing, E., Salim, A., Wang, T., Danesh, J., … Dichgans, M. (2019). Genomic risk score offers predictive performance comparable to clinical risk factors for ischaemic stroke. Nature Communications, 10(1). 10.1038/s41467-019-13848-1

18. Swinnen, J. V., & Dehairs, J. (2022). A beginner’s guide to lipidomics. Biochemist, 44(1), 20–24. 10.1042/bio_2021_181

19. Detrano, R., Guerci, A. D., Carr, J. J., Bild, D. E., Burke, G., Folsom, A. R., … Kronmal, R. A. (2008). Coronary Calcium as a Predictor of Coronary Events in Four Racial or Ethnic Groups. New England Journal of Medicine, 358(13), 1336–1345. 10.1056/nejmoa072100

20. Arnett, D. K., Blumenthal, R. S., Albert, M. A., Buroker, A. B., Goldberger, Z. D., Hahn, E. J., … Ziaeian, B. (2019, September 10). 2019 ACC/AHA Guideline on the Primary Prevention of Cardiovascular Disease: Executive Summary: A Report of the American College of Cardiology/American Heart Association Task Force on Clinical Practice Guidelines. Circulation. NLM (Medline). 10.1161/CIR.0000000000000677

21. Nieminen, T., Lehtinen, R., Viik, J., Lehtimäki, T., Niemelä, K., Nikus, K., … Kähönen, M. (2006). The Finnish Cardiovascular Study (FINCAVAS): Characterising patients with high risk of cardiovascular morbidity and mortality. BMC Cardiovascular Disorders, 6. 10.1186/1471-2261-6-9

22. Blanco-Gómez, A., Castillo-Lluva, S., del Mar Sáez-Freire, M., Hontecillas-Prieto, L., Mao, J. H., Castellanos-Martín, A., & Pérez-Losada, J. (2016). Missing heritability of complex diseases: Enlightenment by genetic variants from intermediate phenotypes. BioEssays, 38(7), 664–673. 10.1002/bies.201600084

23. Greenland, P., Blaha, M. J., Budoff, M. J., Erbel, R., & Watson, K. E. (2018, July 24). Coronary Calcium Score and Cardiovascular Risk. Journal of the American College of Cardiology. Elsevier USA. 10.1016/j.jacc.2018.05.027

24. Cribari-Neto, F., & Zeileis, A. (2010). Beta regression in R. Journal of Statistical Software, 34(2), 1–24. 10.18637/jss.v034.i02

